# Boosting vaccination uptake using wastewater surveillance: a county-randomized controlled trial

**DOI:** 10.64898/2026.09.14.26362268

**Authors:** Milagros Neyra Blatz, Dana Neigel, David A. Larsen, Brittany L. Kmush

## Abstract

Vaccine hesitancy remains a major public health challenge, highlighting the need for effective strategies to boost vaccine uptake. We conducted a county-randomized trial to evaluate whether a social media campaign grounded in alerts driven by community-level wastewater surveillance could increase COVID-19 and influenza vaccination. Forty New York State counties were selected for an 11-week campaign during fall 2024, with 20 counties assigned to the intervention and 20 serving as controls. The campaign reached more than 16.6 million impressions, with over 77K clicks on the call-to-action link, demonstrating substantial public engagement. During the campaign period, intervention counties administered significantly more COVID-19 vaccine doses (β = 76.7, *P* = 0.01) but fewer influenza vaccine doses (β= −140.9, *P* = 0.04) per 100,000 residents per week than control counties. However, season-level analysis showed no significant differences in COVID-19 or influenza vaccination uptake between groups. These findings highlight the potential of alerts from community-level wastewater surveillance to promote vaccination.

**Trial registration:** # NCT06698497

## Introduction

Vaccines are among the most effective and critical tools for safeguarding public health. The World Health Organization’s Expanded Program for National Immunization (EPI) is estimated to prevent 4-5 million deaths worldwide annually and has the potential to avert an additional 1.5 million deaths with universal global coverage.^1,2^ Despite the enormous protection that vaccines provide, vaccine hesitancy, marked by delays or refusals to vaccinate, is on the rise.^3,4^ Hesitancy toward vaccines reduces overall vaccination rates within communities, heightening the risk of outbreaks of vaccine-preventable diseases like polio and measles, as well as amplifying the impact of seasonal illnesses, such as influenza.^5,6^ The underlying causes of vaccine hesitancy are multifaceted and can vary significantly across demographic groups. Broadly speaking, vaccine hesitancy is often categorized into three primary factors: 1) lack of confidence in effectiveness, safety, or governance of vaccines; 2) perceived low risk of contracting diseases; and 3) issues of availability, accessibility, and convenience of obtaining vaccines.^7,8^

Vaccination rates differ across demographic categories, including race, ethnicity, age, and gender.^9^ Additionally, a marked disparity exists between rural and urban areas, with rural counties consistently showing lower vaccination rates than their urban counterparts both nationally and within New York State.^10,11^ Factors contributing to these discrepancies may include challenges in healthcare access in rural areas, such as limited availability of healthcare providers and insufficient information on how and where to access services.^11–13^

Addressing low vaccination rates is critical to mitigating infectious disease risk at both individual and community levels. Public education and communication about infectious diseases and vaccinations can play a significant role in shifting public perceptions, dispelling widespread misconceptions, and reducing vaccine hesitancy.^8,12^ The effectiveness of health communications campaigns is often determined by the interplay between the message, the materials, and the target communities.^14,15^ Therefore, it is crucial to develop communication strategies that not only advocate for vaccination but are also tailored to the unique needs of the communities that are involved. Many vaccine uptake campaigns utilize the health belief model to identify factors that may encourage individuals to get vaccinated.^16,17^ In the health belief model, a disease’s perceived severity, a person’s perceived susceptibility, and a person’s self-efficacy drive the intention to vaccinate.^18,19^ Within the framework of the Health Belief Model (Fig. 1), we hypothesize that using wastewater surveillance in a social media campaign would amplify an individual’s perceived susceptibility and increase their self-efficacy with a call to action to vaccinate. We test this hypothesis with seasonal vaccines, namely COVID-19 and influenza. Wastewater surveillance is well established for SARS-CoV-2 and has already demonstrated its value as an early indicator of hospitalization trends.^20^ Wastewater surveillance is also useful for influenza, with many wastewater surveillance programs adding influenza to their pathogen profile.^21^ With wastewater surveillance providing a leading indicator of community-level transmission, this study explores whether leveraging a social media communications campaign grounded in wastewater surveillance (perceived susceptibility) can boost vaccine uptake.

**Fig. 1.**
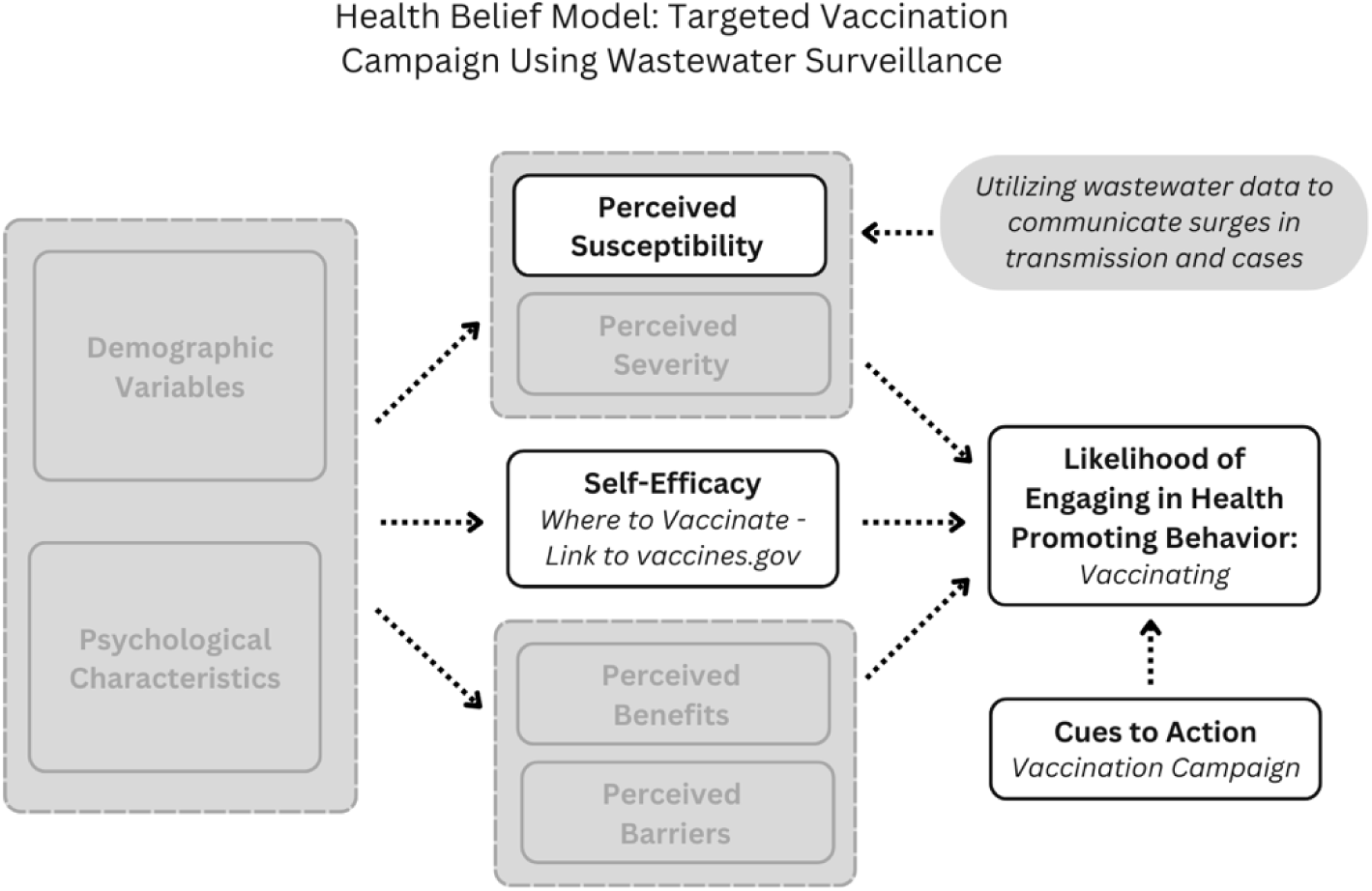
Health Belief Model: Targeted vaccination campaign using wastewater surveillance. The health belief model applied to our hypothesis of using wastewater surveillance to amplify an individual’s perceived susceptibility and increase their self-efficacy. In this model, the cue to action is paired with a direct call to vaccinate. White boxes are facets of the Health Belief Model the campaign is targeting. Grey boxes are facets the campaign does not target.

## Methods

### Setting

New York State is the fourth most populous state in the United States and the 27^th^ state ranked in terms of geographical size. Located in the northeastern United States, much of New York State land, excluding New York City, is rural with a number of upstate urban counties, including Erie (Buffalo, NY), Monroe (Rochester, NY), Onondaga (Syracuse, NY), Oneida (Utica, NY) and Albany (Albany, NY).

Across New York State, uptake of the initial COVID-19 vaccine in 2021 was high, with 90% of the adult population receiving their first series. Subsequent COVID-19 vaccines did not reach these heights; with the 2023-2024 COVID-19 season, vaccine coverage was much lower at 12.3% of the total population (14.7% of adults). Excluding New York City, we selected 40 counties with a combined population of approximately 10,086,086, representing a significant portion (52%) of New York State’s total population for this county randomized trial.^22^ Uptake of the 2023-2024 COVID-19 vaccine in these counties ranged from 6.8% to 23%, with influenza vaccination rates ranging from 17.7% to 34.0% across most study counties, with one substantially higher value (Fig. 2).

**Fig. 2.**
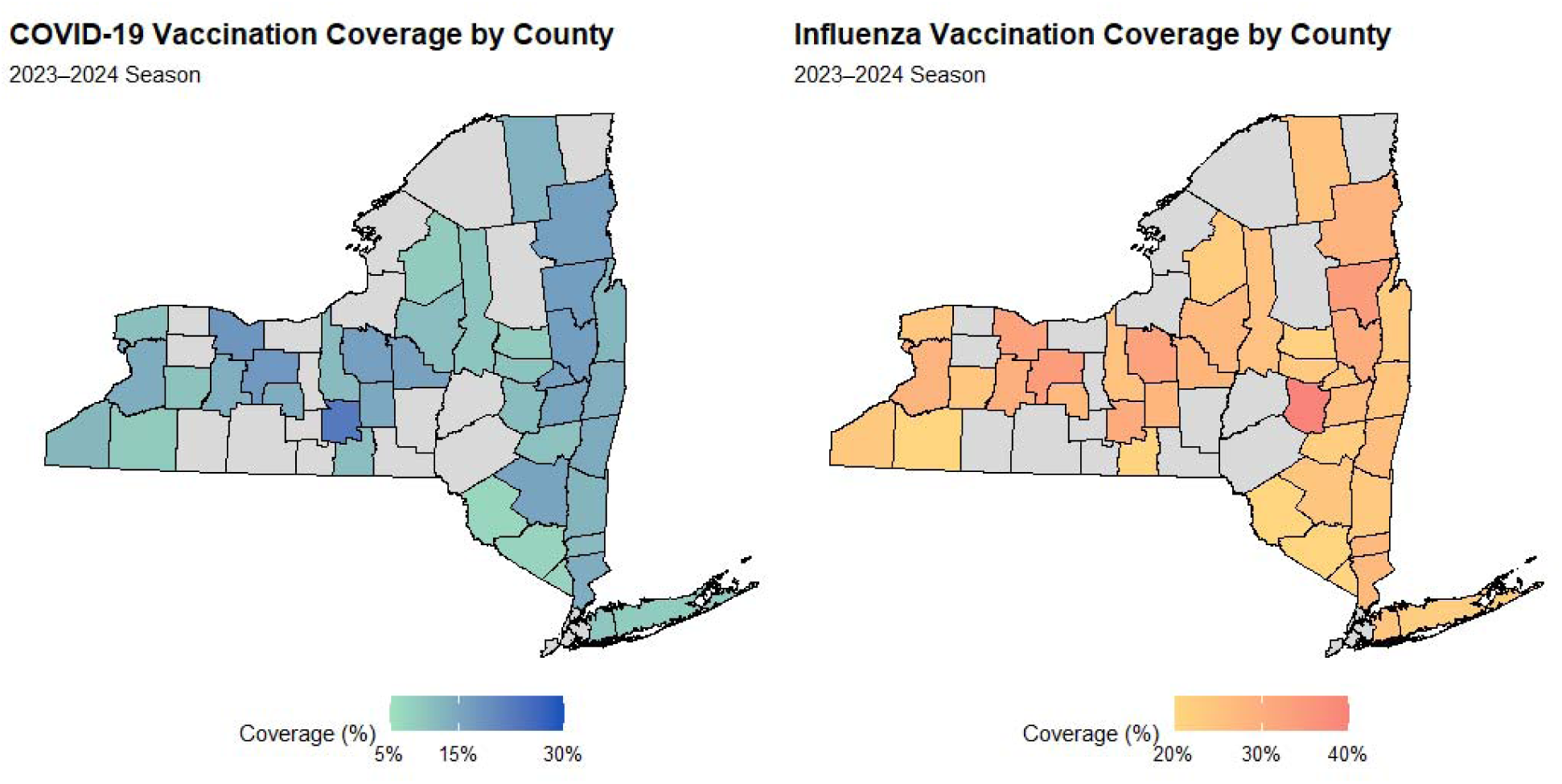
County level vaccine coverage. Seasonal coverage was calculated as total reported 2023-2024 doses administered divided by county population (x100). COVID-19 and influenza color scales were displayed from 5% to 30% and 20% to 40% for visualization clarity, respectively. Gray counties were not included in trial.

### Study Design

This study was designed as a county-randomized controlled trial to assess the effect of a wastewater surveillance-based social media campaign on COVID-19 and influenza vaccine uptake during the fall of 2024 (2024-2025 respiratory virus season). Counties were selected using U.S. Census population data to ensure representation across a range of population sizes and based on their participation in wastewater surveillance. Counties comprising New York City were excluded because their population density was not comparable to the rest of the counties. A total of forty counties were selected for the 11-week campaign in New York State. Selected counties were then randomly assigned to either the control or intervention groups using a lottery-based randomization process, with 20 counties assigned to the intervention group and 20 to the control group. There were no losses or exclusions after randomization. The selected New York State counties in the intervention and control groups are provided in Supplemental Table A. State and local health departments, staff and epidemiologists were blinded to the selection of counties throughout the campaign, with the allocation list held by the research team.

The intervention group received social media advertisements that showcased ongoing wastewater surveillance in their community with messages about getting up to date with their vaccines. The control group received no such advertisements. The target campaign demographic were adults aged 18 and older who were eligible to receive the 2024-2025 COVID-19 and influenza vaccines. Patients or the public were not involved in the design, conduct, or reporting of this research. Campaign adherence and optimization were monitored through OpAD Media, a New York-based media planning and marketing firm.

No adverse events or harms were specifically collected or monitored because the intervention consisted of informational social media advertising and used de-identified, county-aggregated vaccination data. No interim analyses or formal stopping guidelines were specified, as the campaign ran for the prespecified 11-week period. The study protocol was reviewed and deemed exempt by the Institutional Review Board of Syracuse University (IRB #: 23-327). This trial was registered at ClinicalTrials.gov # NCT06698497.

### Communications Campaign

In September 2024, a social media campaign was launched in intervention counties following an increase in SARS-CoV-2 detected in wastewater through the New York State wastewater surveillance network.^23^ Through the network, wastewater samples are routinely analyzed to measure increases or decreases in community-level COVID-19 trends. The data are made publicly available through a dashboard^24^ and shared with county health officials through weekly reports to aid in the understanding of transmission risk.

Campaign advertisements were posted on three social media platforms: Facebook, Instagram, and NextDoor (Fig. 3). Facebook and Instagram creatives featured videos suitable for both desktop and mobile in-feed ads and mobile story ads, while NextDoor creatives used static images for both desktop and mobile in-feed ads. All creatives, including 8 videos and 5 static images, were developed by the research team.

**Fig. 3.**
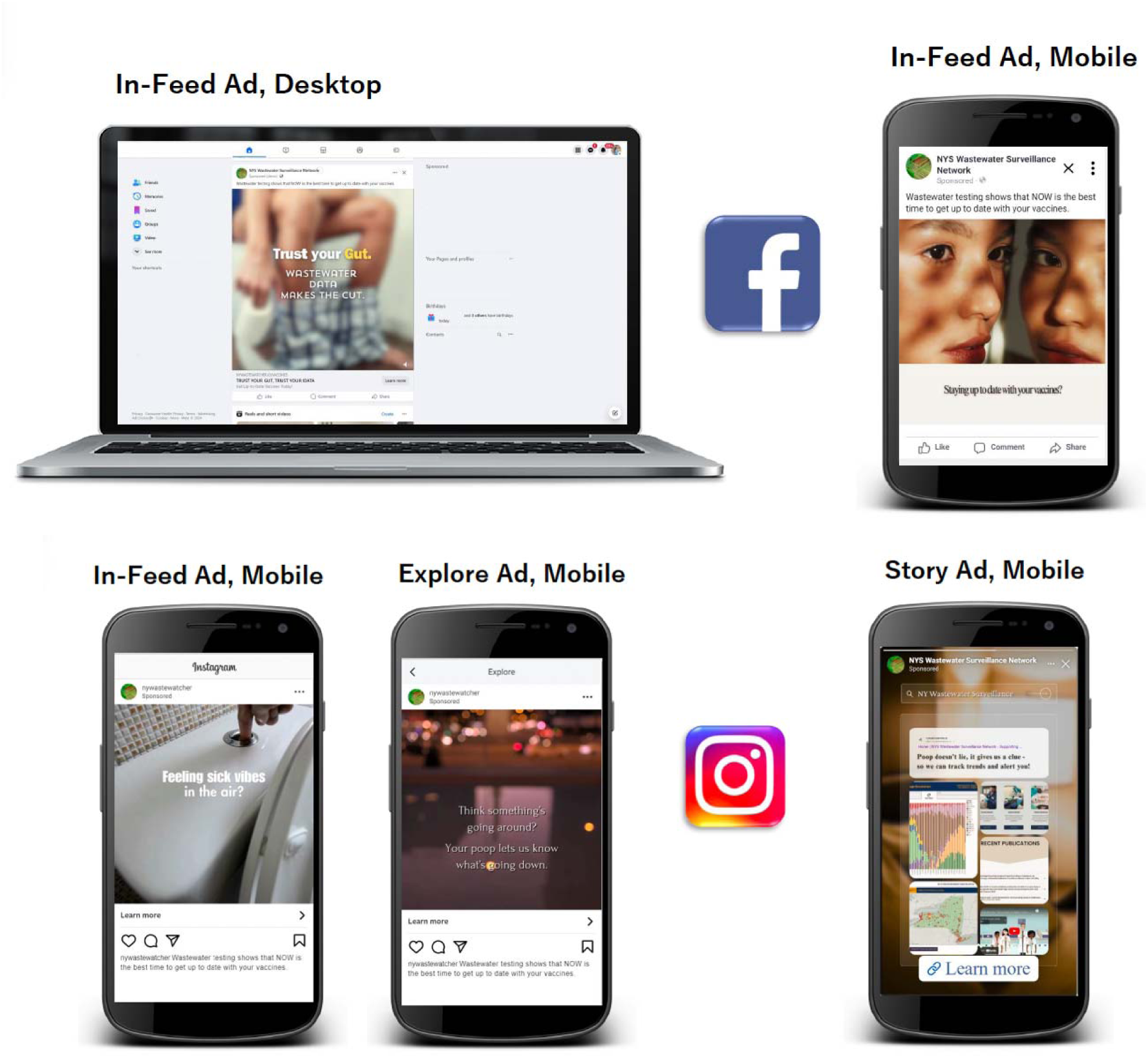
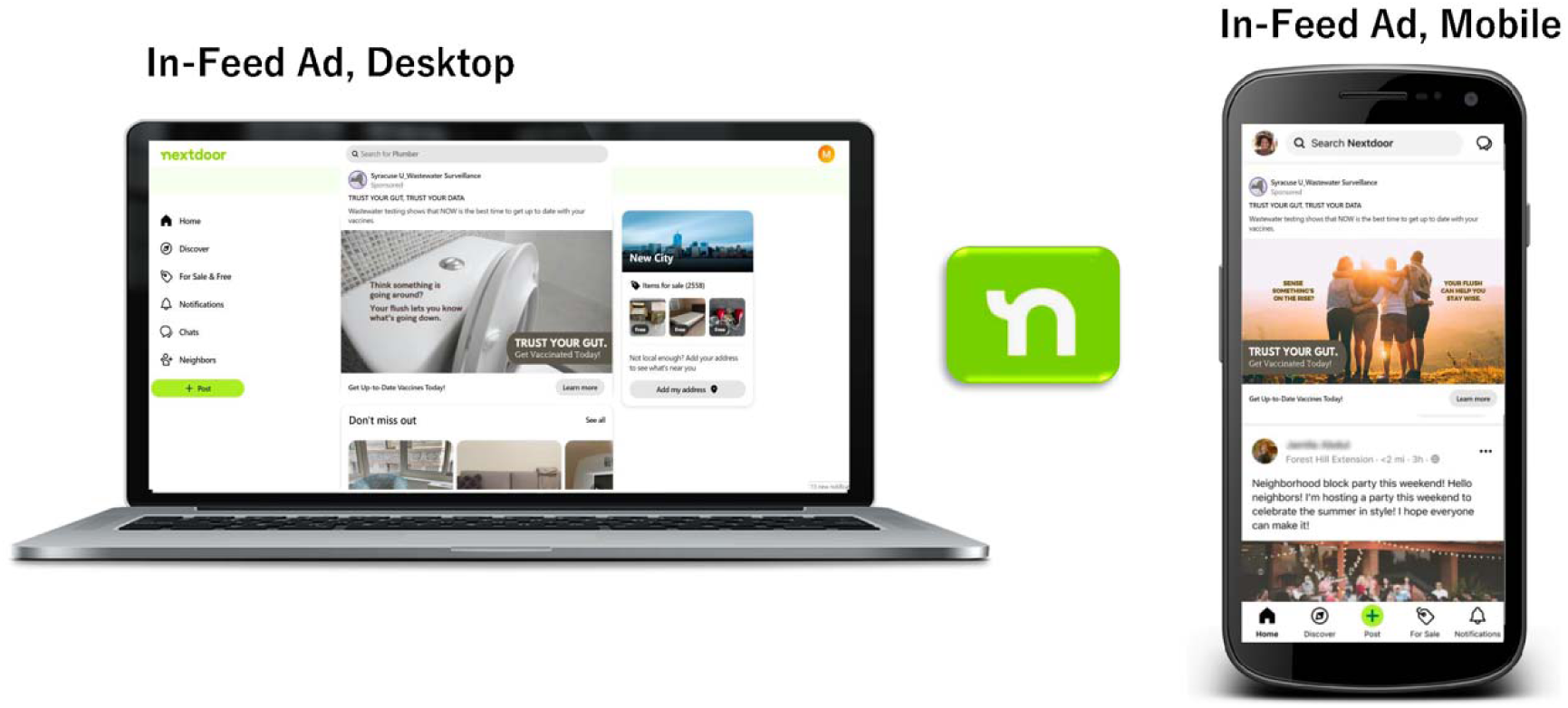
Snapshots of social media creative videos and images. Videos shown in both desktop and mobile in-feed ads for Facebook and Instagram. Images shown in both desktop and mobile in-feed ads for NextDoor. The images shown are mockups of potential ad placements and are not screenshots captured from the live campaign environment. These examples are created by placing the campaign creative within sample platform feeds/templates to demonstrate how the ad would appear in-feed.

The campaign ran for 11 weeks from September 13^th^ through November 24^th^, 2024, and disseminated information highlighting local wastewater surveillance efforts, linking to dynamic COVID-19 wastewater testing results, and encouraging vaccination (not specific to COVID-19 or influenza) among residents in the intervention counties. A substantial portion of the budget was allocated toward targeting residents in zip codes with a high historical vaccination rate for influenza and initial COVID-19 vaccine series. These populations were considered more likely to be receptive to vaccination messages. Prioritizing these areas was intended to maximize campaign reach among individuals most likely to respond to the messaging and seek vaccination within the available budget. Routine NYSDOH vaccination communications and services continued in both groups. All 20 intervention counties received the planned social media campaign; no campaign materials were targeted to control counties.

With expertise from OpAD Media, our social media communications partner, the campaign was continuously optimized across platforms to enhance ad performance metrics. In addition to the creatives (videos and static images), the research team developed complementary social media copy to exemplify the campaign message. The headline read, *“TRUST YOUR GUT, TRUST YOUR DATA,”* paired with the message, *“Wastewater testing shows that NOW is the best time to get up to date with your vaccines.”*

Social media platforms not only offered an efficient medium to disseminate information but also enabled engagement and interaction with community members. Each advertisement included a call-to-action link directing the target audience to a dedicated webpage on the New York State wastewater surveillance network website (nywastewatcher.io/vaccines), which provided information and links regarding vaccines for respiratory-transmitted pathogens (COVID-19 and influenza) as well as guidance on finding vaccine appointment sites.

### Data Sources and Variables

Influenza and COVID-19 vaccination data used in this analysis were obtained from publicly available datasets hosted by the New York State Department of Health (NYSDOH). The data used for this study are routinely collected, quality-checked, and publicly shared by NYSDOH through their open data portal. County-level influenza and COVID-19 vaccination data for the 2024-2025 respiratory virus season was accessed from the New York State COVID-19 and Influenza Vaccination Data portal.^26^ COVID-19 vaccination data for the 2023-2024 season was accessed from the archived New York Statewide COVID-19 Vaccination Data by County (2023-2024) dataset.^25,26^ Influenza vaccination data for the 2023-2024 season were obtained directly from NYSDOH through a formal data request, as these data are not publicly available. These data were provided at the county level and aligned with the publicly available 2024-2025 datasets. Vaccination data are derived from the New York State Immunization Information System (NYSIIS) and provide a lower bound estimate of COVID-19 and influenza vaccination totals.

Additional county-level data were obtained from publicly available sources. Measles, mumps, and rubella (MMR) vaccination coverage was retrieved from the NYSDOH website.^27^ County-level Social Vulnerability Index (SVI) scores were obtained from the New York State Open Data portal.^28^ County metro classification was determined using publicly available documentation and maps from the U.S. Census Bureau and U.S. Department of Agriculture Economic Research Service.^29,30^

All data were publicly accessible and aggregated at the county level. No individual-level or private health information was used or collected as part of this study.

### Statistical Analysis

Weekly county-level dose counts were converted to per-100,000 rates using population denominators and assigned to the appropriate respiratory season. Age categories in the influenza dataset were collapsed to generate total weekly county-level doses. Seasonal cumulative vaccination rates were calculated for each county. For descriptive comparison across seasons, cumulative per-100,000 values were divided by 1,000 to present percent-like units.

Continuous variables were summarized using medians and interquartile ranges. Between group comparisons of baseline county characteristics were assessed using Wilcoxon rank-sum tests. Between-group differences in vaccination uptake were assessed using Welch two-sample t-tests when both groups met normality assumptions and Wilcoxon rank-sum tests otherwise.

Two difference-in-differences (DiD) approaches were used to evaluate vaccination behavior changes: a weekly-level approach within the 2024-2025 respiratory season and a season-level approach comparing the campaign season (2024-2025) with the previous season (2023-2024). Four mixed-effects DiD models were developed:(1) weekly COVID-19 vaccination uptake during the 2024-2025 respiratory season, and (2) weekly influenza vaccination uptake during the 2024-2025 respiratory season, (3) COVID-19 season to season vaccination uptake comparing the 2023-2024 and 2024-2025 respiratory seasons, (4) influenza season to season vaccination uptake comparing the 2023-2024 and 2024-2025 respiratory seasons.

DiD analyses were conducted using mixed-effects regression models. Weekly campaign-period models used weekly vaccination rates per 100,000 residents as the outcome, while season-level models used county-level cumulative vaccination rates per 100,000 residents as the outcome. All models included metropolitan status, county-level SVI, and MMR vaccination coverage as covariates. SVI and MMR were centered, and a county-level random intercept was included to account for repeated observations. The primary parameter of interest was the interaction between intervention group and campaign period (for weekly level models) or season (for season-level models), which tested whether vaccination rate changes differed between intervention and control counties. Difference-in-differences estimates were obtained using mixed-effects regression models, with two-sided p < 0.05 considered statistically significant.

We conducted a simulation-based post-hoc power analysis using the fitted season-level mixed-effects DiD model. Simulations were conducted with a fixed random seed to ensure reproducibility. The model included a county level random intercept and fixed effects for group, season, metro status, centered SVI, and centered MMR coverage. Fixed effect estimates and variance components were extracted from this model.

We simulated datasets across total sample sizes of 40-120 counties and imposed DiD interaction effects ranging from 0-800 additional cumulative doses per 100,000. For each scenario, 1,000 datasets were generated and refit using the same model specification. Power was defined as the proportion of simulations in which the group by season interaction term was statistically significant (p <0.05).

County level covariates (population, SVI, MMR coverage, and metropolitan status) were merged into the analytic datasets prior to modelling. Analyses were restricted to counties included in the randomized trial. Weekly vaccination data contributed to seasonal cumulative rates for all available reporting weeks. All analyses were conducted in R (version 4.2.1).

## Results

### Campaign Performance

The 11-week *Trust your Gut, Trust your Data* campaign reached 16,663,176 million impressions, with 273,617 video completions, and 77,765 clicks on the call-to-action link demonstrating strong public engagement among residents in the 20 intervention counties. Specifically, Facebook and Instagram achieved a unique reach of nearly 2.2 million people, representing about 63% of the target population of approximately 3.5 million across the selected counties. Additionally, the campaign drove a total of 64,352 visits to our website throughout the 11-week campaign duration. Overall, campaign performance metrics demonstrated high fidelity of implementation and achieved broad target population reach (Supplemental Table B).

The overall click-through rate, or the percentage of impressions that result in users clicking the ad, was 0.47%, which is above the campaign benchmark of 0.09% as well as above those reported in similar digital health campaigns.^31^ These results reflect the continuous campaign optimization efforts, which included pausing underperforming creatives and reallocating the budget toward higher-performing social media platforms and ads.

Both reach ads (with the goal of reaching the highest percentage of target audience) and traffic ads (with the goal of driving traffic to the call-to-action landing page) ran across the three platforms. Reach ads achieved a more efficient cost per 1,000 impressions of $3.61, while traffic ads outperformed at a click-through rate of 1.56%. Both performed better than the campaign benchmarks of $3.99 and 1.32%, respectively. Facebook and Instagram drove the more efficient cost per 1,000 impressions ($7.11), whereas NextDoor delivered the higher click-through rate (0.49%).

### Baseline Characteristics

Control and intervention counties were similar across all baseline measures (Table 1). Median population sizes did not differ (88,852 vs 90,608; *P* = 0.72), nor did SVI values (0.418 vs 0.402; *P* = 0.86). MMR vaccination coverage (83.05% and 83.40%; *P* = 0.86) and 2021 COVID-19 vaccination coverage (63.2% vs 60.5%; *P* = 0.48) were also comparable.

**Table 1.** Baseline characteristics of study counties by randomization arm.

| Characteristic | Control* | Intervention* | *P* ** |
| --- | --- | --- | --- |
| Population | 88,900 (57,600 to 338,300) | 90,600 (51,900 to 231,300) | 0.718 |
| Social Vulnerability Index (SVI) | 0.42 (0.25 to 0.63) | 0.40 (0.14 to 0.72) | 0.862 |
| MMR coverage (%) | 83.1 (79.5 to 86.3) | 83.4 (80.1 to 85.5) | 0.860 |
| COVID-19 vaccination coverage, 2021 (%) | 63.2 (57.7 to 68.1) | 60.5 (54.9 to 66.3) | 0.478 |

\*Values are presented as median (interquartile range).
\*\*P-values compare intervention and control counties using Wilcoxon rank-sum tests.

Vaccine Uptake during 2024-2025 Season

Following the intervention in the 2024-2025 respiratory season, mean cumulative COVID-19 vaccination coverage was 10.3% in control counties and 11.6% in intervention counties (Table 2), but the difference was not statistically significant (*P* = 0.22). Mean cumulative influenza vaccination coverage was 27.8% in control counties and 25.2% in intervention counties.

**Table 2.** Seasonal vaccination coverage by study arm, from 2023-2024 to 2024-2025.

| Seasonal Vaccination Coverage | Control | Intervention | *P* ** |
| --- | --- | --- | --- |
| COVID-19 |  |  |  |
| 2023-2024 vaccination rate (per 100k)* | 12,129 | 13,281 | 0.282 |
| 2024-2025 vaccination rate (per 100k)* | 10,331 | 11,567 | 0.217 |
| Percent change (%) | -14.8 | -12.9 | 0.953 |
| **Influenza** |  |  |  |
| 2023-2024 vaccination rate (per 100k)* | 29,458 | 26,331 | 0.490 |
| 2024-2025 vaccination rate (per 100k)* | 27,823 | 25,173 | 0.409 |
| Percent change (%) | -5.6 | -4.4 | 0.947 |

\*Vaccination rates represent mean cumulative doses administered per 100,000 residents during each respiratory season.
\*\*P-values for the 2023-2024 and 2024-2025 rows compare intervention and control counties within season. The p-value for percent change corresponds to the unadjusted difference-in-differences interaction term.

Coverage did not differ significantly between control and intervention counties (*P* = 0.41). Overall, influenza vaccination coverage was higher than COVID-19 vaccination coverage during the 2024-2025 season. However, neither vaccine demonstrated a statistically significant difference in cumulative seasonal coverage between intervention and control counties (Fig. 4).

**Fig. 4.**
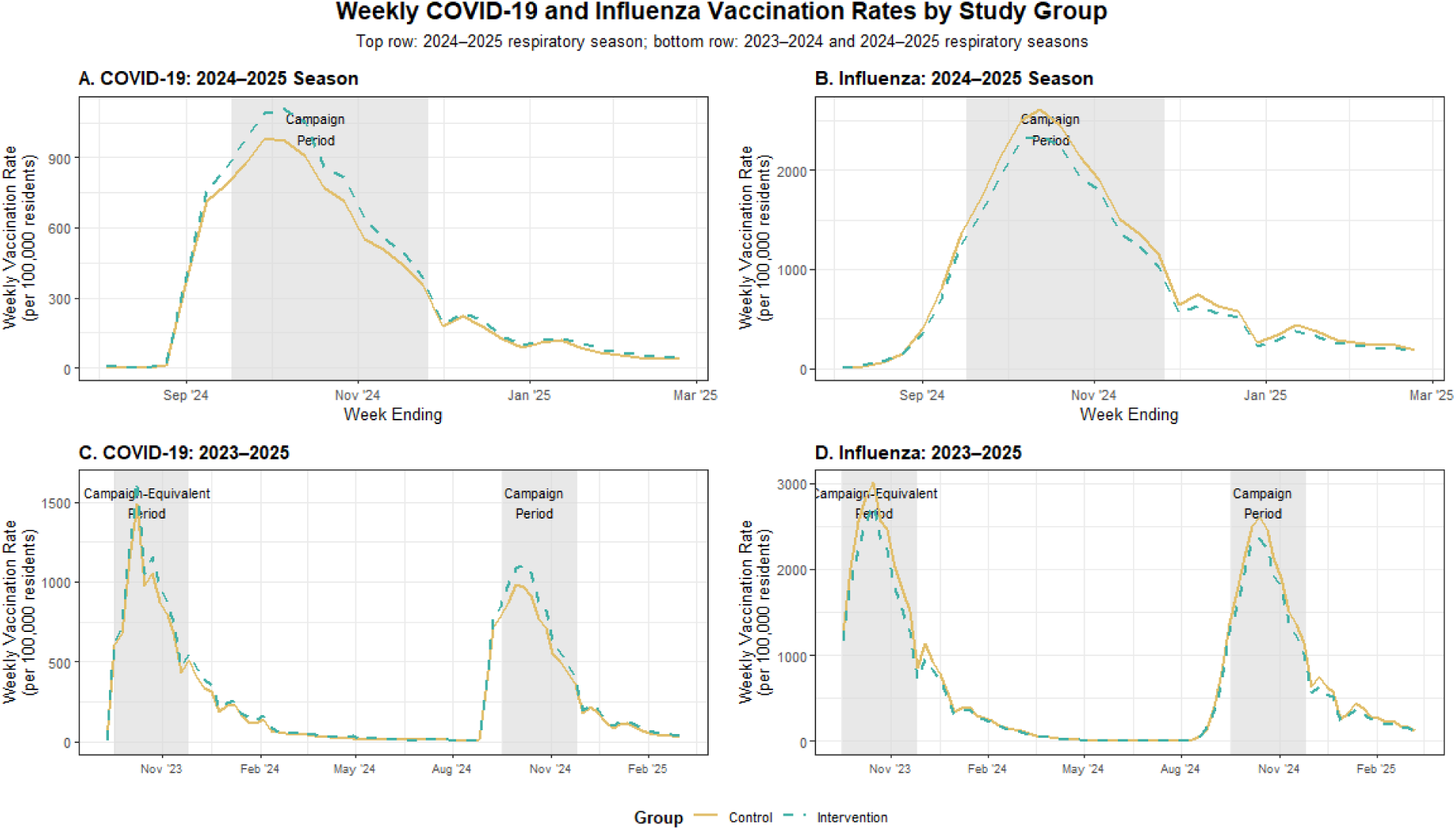
Weekly COVID-19 and influenza vaccination rates per 100,000 residents by study group. Panels **A** and **B** show weekly COVID-19 and influenza vaccination rates, respectively, during the 2024-2025 respiratory season. Panels **C** and **D** show weekly COVID-19 and influenza vaccination rates, respectively, across the 2023-2024 and 2024-2025 respiratory seasons. Shaded areas indicate the campaign period from September 13 through November 24, 2024, and the corresponding calendar period during the preceding season. Lines represent mean county-level weekly vaccination rates for control and intervention counties.

Difference-in-Differences Results

### COVID-19

The weekly-level model examined the 2024-2025 campaign window. When adjusting for the previous 2023-2024 season, intervention counties did not exhibit a meaningfully different year-to-year change in cumulative COVID-19 vaccination uptake compared with control counties (β = 76.7, *P* = 0.01, Table 3, Figure 4 Panel A). In the separate season-level model, the group by season interaction was not statistically significant (β = 85.5, *P* = 0.58), indicating that intervention counties did not exhibit a meaningfully different year-to-year change in cumulative COVID-19 vaccination uptake compared with control counties (Table 3, Figure 4 Panel C).

**Table 3.** Adjusted Difference-in-Differences models of COVID-19 vaccination uptake*.

| Term | Estimate | 95% CI | P |
| --- | --- | --- | --- |
| **A. Weekly COVID-19 vaccination rates before and during the 2024-2025 campaign period** |  |  |  |
| Intercept (control counties, pre-campaign, non-metro) | 137.6 | 74.9 to 200.4 | <0.001 |
| Intervention vs control (pre-campaign) | 11 | -52.1 to 74.1 | 0.727 |
| Campaign period (control counties) | 551.4 | 508.7 to 594.2 | <0.001 |
| Intervention x campaign period (DiD) | 76.7 | 16.3 to 137.1 | 0.013 |
| **Adjustment covariates** |  |  |  |
| Metro vs non-metro | 30.5 | -36.6 to 97.6 | 0.362 |
| Social Vulnerability Index (per 1-point change) | -0.6 | -121.3 to 120.2 | 0.992 |
| MMR coverage (% per 1-point change) | 4.8 | 0.1 to 9.5 | 0.044 |
| **B. Seasonal COVID-19 vaccination rates (2023-2025)** |  |  |  |
| Intercept (control counties, 2023-2024 season, non-metro) | 11,526.1 | 9,566.1 to 13,486.2 | <0.001 |
| Intervention vs control (2023-2024) | 1,018.4 | -910.1 to 2,947.0 | 0.291 |
| 2024-2025 season (control counties) | -1,798.7 | -2,020.1 to -1,577.2 | <0.001 |
| Intervention x 2024-2025 season (DiD) | 85.5 | -227.7 to 398.7 | 0.584 |
| **Adjustment covariates** |  |  |  |
| Metro vs non-metro | 1,030.2 | -1,110.0 to 3,170.5 | 0.335 |
| Social Vulnerability Index (per 1-point change) | -78.9 | -3,932.3 to 3,774.5 | 0.967 |
| MMR coverage (% per 1-point change) | 153.8 | 4.5 to 303.0 | 0.044 |

\*Models were estimated using linear mixed-effects regression with a random intercept for county. Weekly models examined COVID-19 vaccination doses per 100,000 residents across the 2024-2025 respiratory season and compared pre-campaign and campaign periods (September 17-November 26, 2024). Seasonal models examined cumulative COVID-19 vaccination doses per 100,000 residents across the 2023-2024 and 2024-2025 respiratory seasons. All models were adjusted for metropolitan status, centered Social Vulnerability Index (SVI), and centered measles-mumps-rubella (MMR) vaccination coverage. Weekly models included 1,240 county-week observations, and seasonal models included 80 county-season observations across 40 counties.

Overall vaccination levels were lower in 2024-2025 compared with 2023-2024 (β = −1798.7, *P* < 0.001). Among the covariates, only MMR coverage was significantly associated with vaccination uptake (β = 153.8, *P* = 0.04).

### Influenza

The weekly-level model examined the 2024-2025 campaign window. During the campaign period, influenza vaccination increased substantially among control counties (β= 1,672.4, *P* < 0.001), while intervention counties experienced a smaller increase than control counties (β= - 140.9, *P* = 0.035, Table 4, Figure 4 Panel B). This corresponded to approximately 1,532 additional influenza vaccine doses per 100,000 residents per week in intervention counties compared with 1,672 additional doses per 100,000 residents per week in control counties. In the season-level model, the group by season interaction was not statistically significant (β = 478.1, *P* = 0.20), indicating that intervention counties did not exhibit a meaningfully different year-to-year change in cumulative influenza vaccination uptake compared with control counties (Table 4, Figure 4 Panel D). Overall vaccination levels were lower in 2024-2025 compared with 2023-2024 (β= −1,635.5, *P* < 0.001). None of the covariates were significantly associated with influenza vaccination uptake.

**Table 4.** Adjusted Difference-in-Differences models of Influenza vaccination uptake*.

| Term | Estimate | 95% CI | P |
| --- | --- | --- | --- |
| **A. Weekly Influenza vaccination rates before and during the 2024-2025 campaign period** |  |  |  |
| Intercept (control counties, pre-campaign, non-metro) | 188.8 | -72.5 to 450.1 | 0.151 |
| Intervention vs control (pre-campaign) | -32.4 | -290.1 to 225.4 | 0.800 |
| Campaign period (control counties) | 1,672.4 | 1,579.9 to 1,764.9 | <0.001 |
| Intervention x campaign period (DiD) | -140.9 | -271.7 to -10.2 | 0.035 |
| **Adjustment covariates** |  |  |  |
| Metro vs non-metro | 137.5 | -147.2 to 422.2 | 0.334 |
| Social Vulnerability Index (per 1-point change) | -217.9 | -730.5 to 294.7 | 0.394 |
| MMR coverage (% per 1-point change) | -2 | -21.8 to 17.9 | 0.843 |
| **B. Seasonal Influenza vaccination rates (2023-2025)** |  |  |  |
| Intercept (control counties, 2023-2024 season, non-metro) | 25,944.6 | 15,354.2 to 36,535.0 | <0.001 |
| Intervention vs control (2023-2024) | -3,186.5 | -13,594.6 to 7,221.6 | 0.538 |
| 2024-2025 season (control counties) | -1,635.5 | -2,161.7 to -1,109.3 | <0.001 |
| Intervention x 2024-2025 season (DiD) | 478.1 | -266.1 to 1,222.3 | 0.201 |
| **Adjustment covariates** |  |  |  |
| Metro vs non-metro | 5,451.1 | -6,126.0 to 17,028.2 | 0.346 |
| Social Vulnerability Index (per 1-point change) | -8,724.4 | -29,568.4 to 12,119.5 | 0.401 |
| MMR coverage (% per 1-point change) | -73.2 | -880.6 to 734.1 | 0.855 |

\*Models were estimated using linear mixed-effects regression with a random intercept for county. Weekly models examined influenza vaccination doses per 100,000 residents across the 2024-2025 respiratory season and compared pre-campaign and campaign periods (September 13-November 26, 2024). Seasonal models examined cumulative influenza vaccination doses per 100,000 residents across the 2023-2024 and 2024-2025 respiratory seasons. All models were adjusted for metropolitan status,

### Post hoc power analysis

For the season-level COVID-19 difference-in-differences model, the empirical Type I error under a null intervention effect was 0.04. With 40 counties, power was 0.24, 0.45, and 0.88 for DiD effects of 200, 300, and 500 additional cumulative doses per 100,000 residents, respectively, indicating that the current study design was adequately powered to detect large effects but underpowered to detect more modest ones (Table 5).

**Table 5.** Empirical type I error and post hoc power estimates from simulation*.

| Number of Counties | Type I Error | $\beta = 100$ | $\beta = 200$ | $\beta = 300$ | $\beta = 500$ | $\beta = 800$ |
| --- | --- | --- | --- | --- | --- | --- |
| 40 | 0.04 | 0.10 | 0.24 | 0.45 | 0.88 | 1.00 |
| 60 | 0.04 | 0.11 | 0.35 | 0.63 | 0.98 | 1.00 |
| 80 | 0.07 | 0.15 | 0.43 | 0.77 | 0.99 | 1.00 |
| 100 | 0.05 | 0.19 | 0.53 | 0.87 | 1.00 | 1.00 |
| 120 | 0.05 | 0.19 | 0.58 | 0.91 | 1.00 | 1.00 |

\*Power estimates are based on 1,000 simulated datasets for each combination of county sample size and difference-in-differences (DiD) effect. The DiD effect represents additional cumulative vaccine doses per 100,000 residents attributable to the intervention.

## Discussion

Here we show mixed evidence of the effect of a social media campaign driven by localized wastewater surveillance data to increase seasonal vaccination uptake. Improving a public health surveillance system, including continuous wastewater surveillance for infectious disease, increases the capacity to implement public health interventions more precisely.^32^ Wastewater surveillance offers geographically based and locally relevant data about infectious disease risks to communities. We hypothesized that this more localized information provided by wastewater surveillance would improve seasonal respiratory virus vaccine uptake. This theory of local information having an impact on health behavior is a novel application of the health belief model.

Previous vaccination campaigns based on the health belief model have highlighted the perceived benefits, perceived barriers, and cues to action as the most dominant constructs driving intention to vaccinate. In addition, perceived susceptibility had a stronger influence on vaccination intention than perceived severity.^33^ Our campaign to promote vaccination uptake focused on the Health Belief Model constructs of cues to action, self-efficacy, and perceived susceptibility, which is enabled by enhanced infectious disease surveillance in wastewater. Our application of perceived susceptibility differs from other vaccination campaigns in that the risk information shared in the campaign was geographically relevant at a local community level and responsive to changes in respiratory virus activity detected in wastewater. Rather than communicating susceptibility as a static risk, the campaign used messaging based on data from New York State’s wastewater surveillance dashboard and provided links for users to directly view viral activity in their communities. This localized information intended to make susceptibility feel more personally relevant. The social media advertisements also served as a cue to action, while our links to vaccination scheduling and site locations provided an actionable next step intended to support self-efficacy.

We found that during the campaign period, COVID-19 vaccination was increased by approximately 77 additional weekly doses per 100,000 residents in the intervention counties compared to the control counties. However, this pattern did not hold true for influenza vaccination or when we compared overall seasonal vaccination from the previous year. Although weekly COVID-19 uptake was higher in intervention counties during the campaign, the absence of a significant season-level effect suggests that this increase did not translate into a measurable difference in cumulative seasonal vaccination. Likewise, although influenza vaccination increased substantially during the campaign period overall, intervention counties experienced a smaller weekly increase than control counties, and this difference was not reflected in cumulative seasonal vaccination.

Our findings are similar to previous studies to increase vaccine uptake through incentives, improving access, education, and communication. A literature review of 235 interventions evaluated in 144 randomized trials found that 35% of trials showed no significant effect of the intervention on vaccine uptake, with effect sizes ranging from −6 to 50 percentage points.^34^ Even interventions offering financial incentives have not consistently increased vaccination uptake.

State vaccine lotteries were associated with a 23.1% relative increase in the daily first dose vaccination rate, corresponding to about 53 additional doses per 100,000 residents per day.^35^ However, results varied substantially across states, with effects not being observed in several. More intensive vaccination uptake strategies that directly address access barriers have sometimes produced notable changes, with mobile vaccine delivery in Sierra Leone increasing uptake by 26 percentage points in remote communities.^34^ Our campaign differed from both approaches. Rather than offering financial incentives or directly bringing vaccinations to communities, we used localized wastewater information and social media messaging to encourage vaccination uptake. With this in mind, the approximate 77 additional weekly COVID-19 doses per 100,000 residents observed during the campaign shows a modest short-term bump in vaccination uptake. However, the absence of a cumulative seasonal COVID-19 and influenza effect indicates that the campaign did not consistently increase uptake across outcomes. Although differences in setting, vaccines, and outcomes prevent a direct comparison to other noted interventions, these findings are consistent with the variability in effectiveness across vaccination uptake interventions.

Changing vaccination behavior is challenging, and perceived risk is only one of many factors that influence uptake. Attitudes towards vaccines, perceived benefits and barriers, social norms, and practical considerations such as access and convenience can also affect whether an individual receives a vaccine.^4,6,8^ Given the complexity of vaccination decision making, an increased awareness of local disease activity may not necessarily translate into vaccine uptake. We did not measure vaccine attitudes or intentions, perceived susceptibility, or self-efficacy, so we are unable to determine whether the campaign influenced an individual’s underlying beliefs and intentions.

The study also presented an opportunity to observe the cost-effectiveness of a multimedia campaign informed by infectious disease surveillance. It is generally understood that infectious disease surveillance has value, particularly in identifying emerging threats. Unfortunately, the standard approach to assessing the cost-effectiveness of investment in public health interventions through cases, hospitalizations and deaths prevented is difficult to apply to infectious disease surveillance.^36^ By linking the digital intervention to the signal from wastewater surveillance, researchers may infer how wastewater surveillance might increase COVID-19 vaccination uptake and therefore estimate a cost-effectiveness of wastewater surveillance to prevent COVID-19 hospitalizations.

### Limitations and future directions

This study has several limitations. Because vaccination outcomes were assessed at the county level, individual exposure to or engagement with campaign materials could not be linked to vaccination intentions or subsequent vaccination behavior, and uptake may have been influenced by other vaccination efforts or unmeasured county-level factors. The 11-week campaign may have been too brief to produce sustained changes in seasonal vaccination, and the study had limited power to detect modest season-level effects. In addition, the campaign messaging did not specify COVID-19 or influenza vaccination and the New York State wastewater dashboard displayed COVID-19 but not influenza data, making influenza vaccination a less direct test of the intervention. Future studies should evaluate longer or repeated campaigns across multiple respiratory seasons, include larger samples and pathogen specific wastewater data, and examine individual-level relationships between campaign exposure, engagement, and vaccination intentions and behavior.

### Implications for policy

These findings suggest that wastewater surveillance may support public health practice not only by monitoring infectious disease trends, but also by informing timely, geographically precise communication. The campaign performed favorably compared with pre-determined benchmarks, demonstrating that wastewater surveillance-informed messaging can effectively engage the public. The use of “potty” humor, including wastewater-related jokes and toilet imagery, may have helped attract attention and make the information more approachable. The engagement observed could be used to good effect during a public health emergency such as an outbreak of a vaccine-preventable disease. The public health response to the 2022 polio outbreak in New York State was enhanced with understanding from wastewater surveillance,^37,38^ but the vaccine communications campaign was not wastewater themed. Wastewater surveillance-informed communication may be most useful as part of a broader vaccination strategy, while future campaigns should continue to examine how creative messaging can translate public engagement into health behavior action.

## Data Availability

The vaccination data that supports the findings of this study are publicly available in the New York State Department of Health dashboards, https://coronavirus.health.ny.gov/covid-19-and-influenza-vaccination-data and https://health.data.ny.gov/Health/New-York-State-Statewide-COVID-19-Vaccination-Data/gikn-znjh/about_data. Influenza vaccination data for the 2023-2024 season are available with permission of the New York State Department of Health through a formal data request. The campaign performance data are available from the corresponding author upon reasonable request.

## Supplementary material

## Acknowledgements

This study was supported by the Centers for Disease Control and Prevention (CDC) Epidemiology and Laboratory Capacity (ELC) Program, funded through Health Research, Inc (HRI) and the New York State Department of Health (NYSDOH) with Federal Award Identification Number NU50CK000516. The contents are those of the author(s) and do not necessarily represent the official views of, nor an endorsement by, NYSDOH, HRI, or CDC.

We gratefully acknowledge OpAD Media as our communications partner in the implementation of the social media campaign and for providing snapshots of the social media creatives included in this work.

## Author contributions

Conceptualization: D.A.L., M.N.B. Data curation: D.T.H., M.N.B., D.N. Formal analysis: D.N. Funding acquisition: D.A.L., M.N.B. Methodology: M.N.B., D.N. Project administration: D.A.L., M.N.B., B.L.K. Supervision: D.A.L., M.N.B., B.L.K. Visualization: D.A.L., M.N.B., D.N. Writing – original draft: M.N.B., D.N. Writing – review & editing: D.A.L., B.L.K.

---

**Supplemental Table A.** List of New York State Counties included in the intervention and control study arms.

| **NYS Counties in the Intervention Group (N = 20)** | **NYS Counties in the Control Group (N = 20)** |
| --- | --- |
| Cattaraugus | Albany |
| Chautauqua | Cayuga |
| Cortland | Columbia |
| Dutchess | Franklin |
| Erie | Greene |
| Essex | Lewis |
| Fulton | Madison |
| Herkimer | Monroe |
| Livingston | Nassau |
| Montgomery | Niagara |
| Oneida | Orange |
| Onondaga | Putnam |
| Ontario | Rensselaer |
| Rockland | Schoharie |
| Saratoga | Suffolk |
| Schenectady | Sullivan |
| Tioga | Ulster |
| Tompkins | Washington |
| Warren | Westchester |
| Wyoming | Yates |

**Supplemental Table B.** Campaign performance metrics, from September 13, 2024, to November 24, 2024.

| Metric | Value |
| --- | --- |
| **Overall performance** |  |
| Impressions | 16,663,176 |
| Clicks on call-to-action link | 77,765 |
| Click-through rate | 0.47% |
| Video completions | 273,617 |
| Video completion rate | 1.74% |
| Cost per 1,000 impressions | \$7.23 |
| **Performance by platform** |  |
| **Facebook & Instagram:** |  |
| Impressions | 15,745,214 |
| Click-through rate | 0.47% |
| Video completion rate | 1.74% |
| Cost per 1,000 impressions | \$7.11 |
| **NextDoor:** |  |
| Impressions | 917,962 |
| Click-through rate | 0.49% |
| Cost per 1,000 impressions | \$9.24 |
| **Performance by objective** |  |
| **Reach ads:** |  |
| Impressions | 12,227,003 |
| Click-through rate | 0.07% |
| Video completion rate | 0.68% |
| Cost per 1,000 impressions | \$3.61 |
| **Traffic ads:** |  |
| Impressions | 4,436,173 |
| Click-through rate | 1.56% |
| Video completion rate | 4.68% |
| Cost per 1,000 impressions | \$17.22 |

